# Significant Reduction in Manual Annotation Costs in Ultrasound Medical Image Database Construction Through Step by Step Artificial Intelligence Pre-annotation

**DOI:** 10.1101/2025.01.09.25320077

**Authors:** Fu Zheng, Liu XingMing, Xu JuYing, Tao MengYing, Yang BaoJian, Shan Yan, Ye KeWei, Lu ZhiKai, Huang Cheng, Qi KeLan, Chen XiHao, Du WenFei, He Ping, Wang RunYu, Ying Ying, Bu XiaoHui

## Abstract

This study investigates the feasibility of reducing manual image annotation costs in medical image database construction by utilizing a step by step approach where the Artificial Intelligence model(AI model) trained on a previous batch of data automatically pre-annotates the next batch of image data, taking ultrasound image of thyroid nodule annotation as an example. The study used yolov8 as the AI model. During the AI model training, in addition to conventional image augmentation techniques, augmentation methods specifically tailored for ultrasound images were employed to balance the quantity differences between thyroid nodule classes and enhance model training effectiveness. The study found that training the model with augmented data significantly outperformed training with raw images data. When the number of original images number was only 1,360, with 7 thyroid nodule classifications, pre-annotation using the AI model trained on augmented data could save at least 30% of the manual annotation workload for junior physicians. When the scale of original images number reached 6,800, the classification accuracy of the AI model trained on augmented data was consistent with that of junior physicians, eliminating the need for manual preliminary annotation.

## 1. Introduction

The application of artificial intelligence (AI) in the medical field is booming, and AI-assisted medical image classification and recognition are gradually being deeply integrated into clinical practice, widely used in CT, MRI, pathological diagnosis, and ultrasound imaging-aided diagnosis [1-4], becoming a valuable assistant to doctors. AI models require targeted training to perform related tasks. AI model training methods are generally divided into unsupervised learning [5], semi-supervised learning [6], and supervised learning [7] based on whether supervision is present. For medical image classification tasks, the predicting results of supervised learning is more close to classification habits in clinical practice.

Supervised learning requires images to be annotated in advance. Excellent AI models rely on meticulously and professionally annotated medical image data. The quality of image data annotation directly affects model performance [8]. The annotation of ultrasound medical image data often requires preliminary annotation by junior physicians followed by review by senior physicians to ensure the accuracy of annotation information. Therefore, doctors’ experience directly determines the quality of image annotation, and good doctors are scarce and busy with clinical work, resulting in high costs for manual annotation, which limits the scale of image databases. The performance of AI is related to the size of the training data set; thus, how to reduce manual annotation costs and rapidly expand the database scale is a critical issue affecting AI model quality.

Additionally, medical images exhibit imbalanced class distributions, where some rare diseases occurring rates are less then 0.1%, and some common ones are more then 30%, which significantly reduces model training efficiency. Studies have shown that data augmentation can effectively improve model training efficiency [9]. These augmentation methods are widely used for images captured by cameras, but the generation principles of ultrasound image data differ from those of camera-captured images. Therefore, how to specifically augment ultrasound images is an urgent problem to be solved to enhance training effectiveness with a fixed database scale.

Current research indicates that the accuracy of AI models in image annotation and classification can surpass manual annotation [10], making it a feasible solution to let AI models assist in annotation while learning.

This study takes thyroid nodule ultrasound images as an example to explore the feasibility of using AI to pre-annotate images to save manual annotation costs when dealing with small sample sizes and significant quantity differences between classes in ultrasound image data.

## 2. Experimental Methods

### 2.1 Database Image Source and Composition

This study selected 8,500 ultrasound images of various thyroid nodules from the workstation database of the Ultrasound Diagnostic Department of the 906th Hospital. These selected grayscale images do not contain color Doppler blood flow signals, indicator arrows, measurement marks, or other auxiliary marks. Each image contains at least one thyroid nodule. The sections include coronal (short-axis) and sagittal (long-axis) sections of the thyroid gland, as well as non-standard sections. We conducted privacy desensitization, ensuring that the images did not contain patient name information.

### 2.2 Database Image Annotation

We conducted two-stage annotation of ultrasound images. In the first stage, junior physicians (residents, physicians) annotated each image, and in the second stage, physicians above the deputy senior level (associate chief physicians, chief physicians) reviewed and corrected the annotations from the first stage.

We saved the annotation results from junior physicians in the first stage and the review results from senior physicians in the second stage. The model was trained using the annotated data set after the second-stage. We used the review results from senior physicians as the gold standard to calculate the misannotation rate of junior physicians in the first stage.

Various international standards for thyroid nodule classification exist, such as the 2017 ACR TI-RADS (Thyroid Imaging Reporting and Data System) by the American College of Radiology [11], the 2015 TI-RADS standard by the American Thyroid Association (ATA) [12], and the Korean TI-RADS [13]. In China, in addition to these international standards, the TI-RADS standard established by Shangyan Xu, Weiwei Zhan, et al., in 2016 [14], and the 2020 CTI-RAIDS classification [15] are also commonly used. Since our hospital consistently adopts the TI-RADS classification standard established by Shangyan Xu and Weiwei Zhan in 2016[14], and the participating physicians are more familiar with this classification system, this study used this standard to annotate thyroid nodules in the images. This classification standard divides nodules into 2, 3, 4A, 4B, 4C, 5, and 6 categories. Since the 6 category in the classification standard is based on pathological diagnosis, not image morphology, only categories 2-5 are annotated in this study. For example, if an ultrasound image shows a nodule classified as 4B and the Fine Needle Aspiration Biopsy(FNAB) pathology result of this nodule indicates thyroid cancer, it should be annotated as category 6, but our database still annotates such images as category 4B based on their image manifestations. Fig 1 shows the quantity, size, and location distributions of these nodule categories.

**Fig 1:**
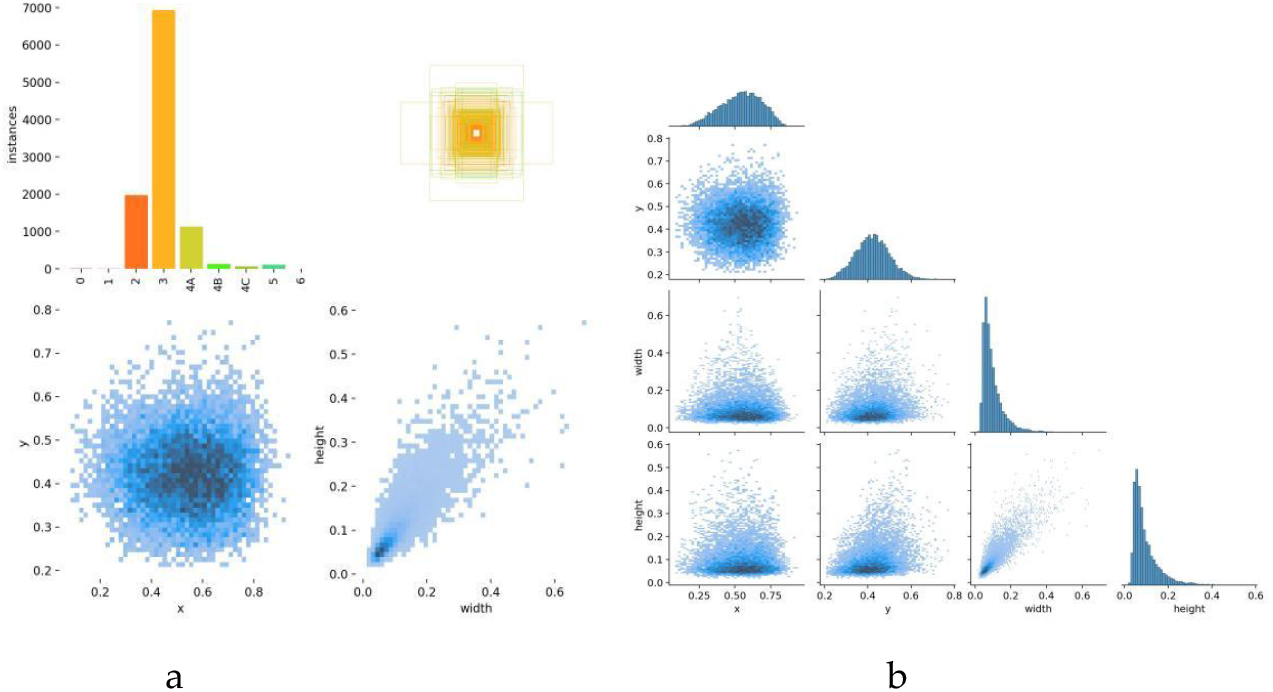
Distribution of nodule categories, sizes, and locations in the image data set. The bar chart in the upper left corner of Fig 1a represents the data quantities of each category in the training set. Each colored bar represents the quantity of nodules in each category. In the upper right, lower left, and lower right corners of Fig 1a respectively show the sizes and quantities of bounding boxes for nodules, the positions of nodule centers relative to the entire image, and the aspect ratios of nodules relative to the entire image. The Fig 1b represents the relationships between the center coordinates x and y, and the width and height of the bounding boxes.

**Fig 2:**
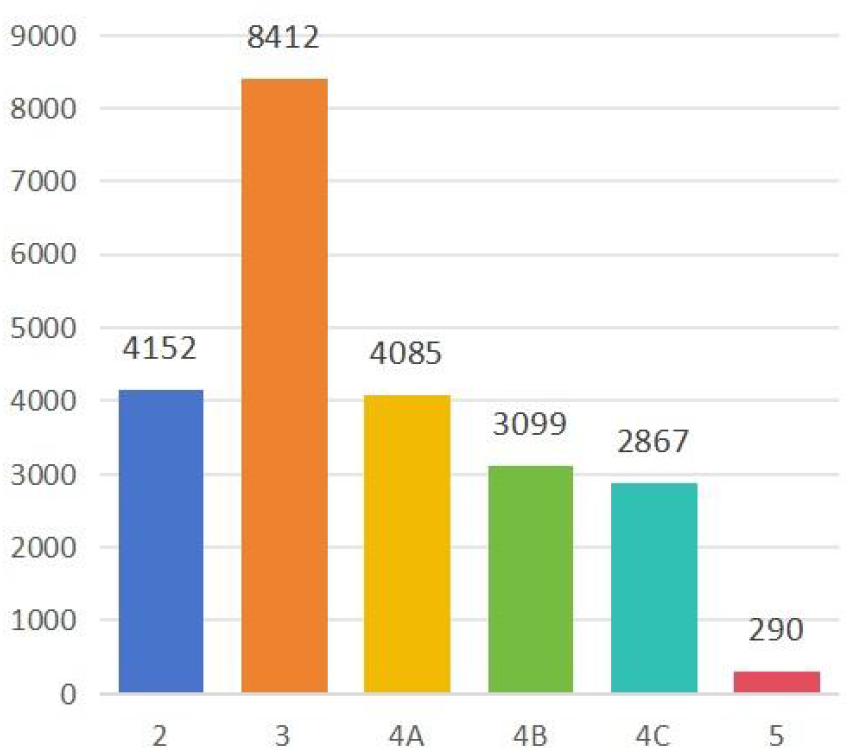
Bar chart of nodule category distributions after image augmentation. Fig 2 shows that although category 3 nodules still have the highest quantity, compared to Fig 1, the quantities of categories 4A, 4B, and 4C have significantly increased, approaching half the quantity of category 3 nodules, and the quantity of category 5 nodules has also increased somewhat. Since multiple nodules may appear in a single image, augmenting images with fewer nodules will also increase the quantity of category 3 nodules to some extent. Therefore, compared to Fig 1, the quantity of category 3 nodules has also increased slightly.

### 2.3 Data Augmentation

Since categories 4A, 4B, 4C, and 5 contain very few nodules, class imbalance may lead to poor training effectiveness [16]. Therefore, it is necessary to augment images containing these rare nodule types to improve AI model training effectiveness. In addition to conventional image augmentation methods such as brightness and contrast changes and small angle rotations [17], we specifically included ultrasound-specific augmentation methods such as defocus, acoustic shadow, and sidelobe artifacts. Studies have shown that using too many augmentation methods on a single image may not enhance training effectiveness [18], so we randomly applied only two augmentation methods to each image.

The ultrasound-specific augmentation methods adopted in this study are as follows:

#### Simulated Defocus

Similar to optical imaging systems, ultrasound imaging systems may also experience out-of-focus situations, resulting in blurred images. We used Gaussian blur to blur the images to a certain extent, simulating defocus. Gaussian blur is a convolution operation where the Gaussian convolution kernel is a two-dimensional matrix whose element values are calculated based on the Gaussian distribution function. This convolution kernel is then convolved with each pixel and its neighboring pixels in the image to obtain new pixel values.

#### Simulated Acoustic Shadow

Similar to how shadows are produced behind opaque objects in optical systems, acoustic shadows are dimmed areas produced when sound beams encounter calcified lesions or other tissues that are difficult for sound beams to penetrate. In this study, we wrote a code to randomly generate dimmed areas of random positions and sizes in the images.

#### Sidelobe Artifacts

Due to the characteristics of ultrasound probes, sidelobes interfere with the main beam, resulting in a relatively faint rotated and displaced original image superimposed on the image. We wrote a code to achieve this effect.

### 2.4 AI Model Selection

This study used the yolov8 model for training. YOLOv8 is a product of the YOLO architecture model family from Ultralytics[19], capable of object detection, instance segmentation, and image classification. This model, created in PyTorch[20], can run on both CPUs and GPUs. YOLOv8 is an accurate and flexible visual AI solution that excels in various visual technology tasks and demonstrates superiority in multiple aspects. YOLO means “You Only Look Once,” as it can annotate target types and coordinates throughout an image with a single scan. It is widely used in image classification, segmentation, and other fields.

### 2.5 AI Model Training

We trained the model in two stages: once without data augmentation and once with data augmentation.

#### 2.5.1 AI Model Parameter Settings

**Table 1:**
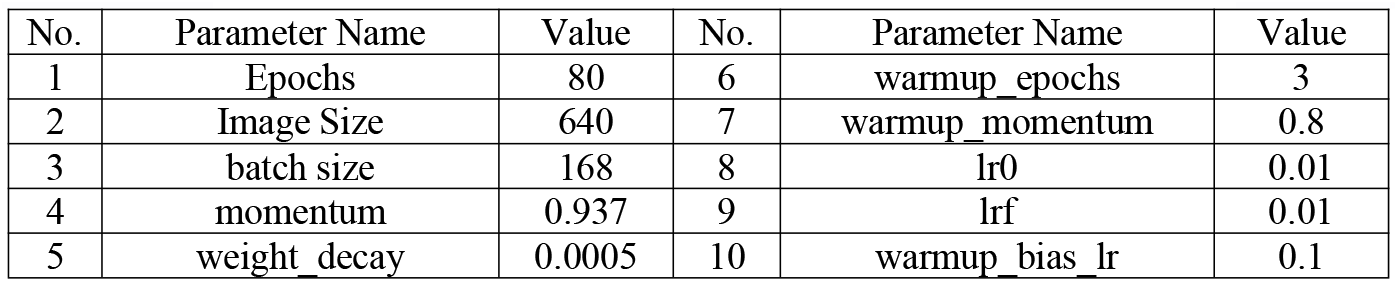
Main parameters for AI model training.

**Table 2:**
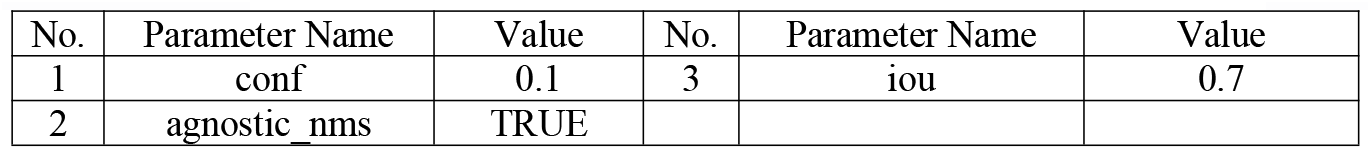
Main parameters for image AI model prediction. Both model training and image prediction utilized the NVIDIA RTX 4090 GPU for computational acceleration.

#### 2.5.2 Model Training Without Data Augmentation

We first trained the model without data augmentation to compare its effectiveness with training using augmented data. The data set was equally divided into 25 portions, and model training was conducted in 5 batches. The Nth batch used the first 5×N portions of data in the database, with yellow portions serving as the training data set and blue portions as the validation data set (see Fig 3). For example, in the second batch, we used the first 5×2=10 portions of data for training, with 8 yellow portions for the training data set and 2 blue portions for the validation data set.

**Fig 3:**
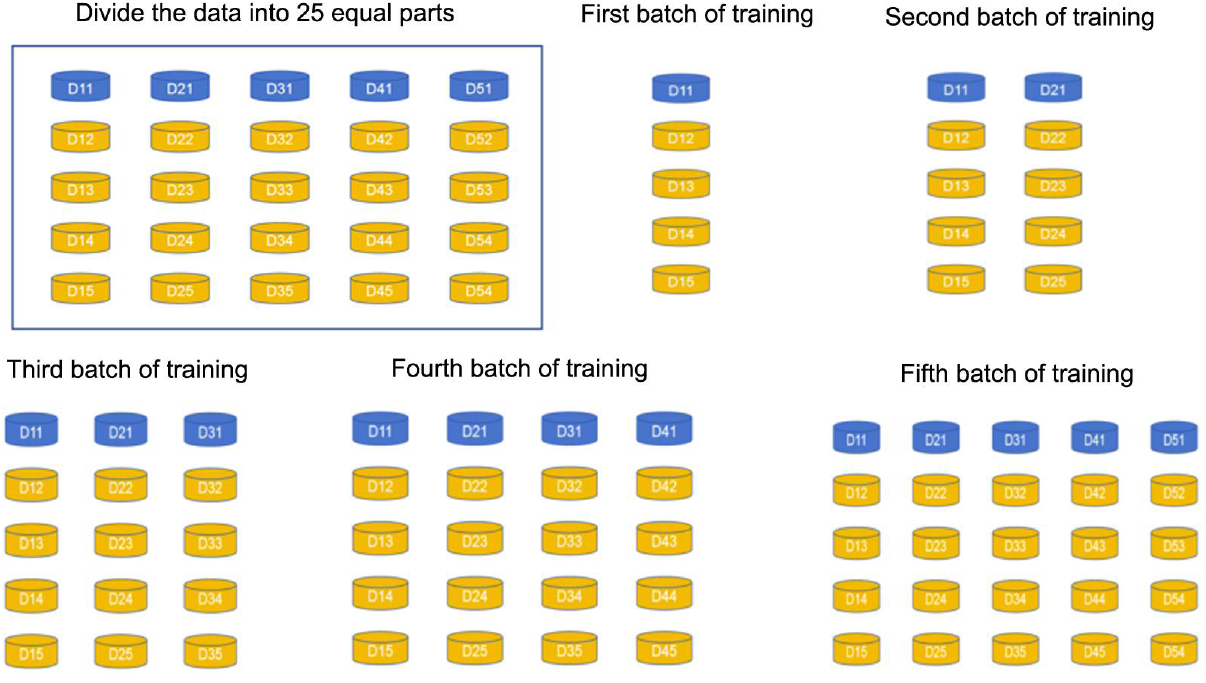
Data division for each batch of training. Fig 3 shows that the entire database was randomly divided into 25 equal portions and trained in 5 batches. The Nth batch used the first 5×N portions of data as the database, with yellow portions serving as the training data set and blue portions as the validation data set. For example, in the second batch, we used the first 10 portions of data for training, with 8 yellow portions for the training data set and 2 blue portions for the validation data set.

Each batch underwent 80 epochs of training. The network weight parameters of the first batch’s training model were randomly initialized, and after 80 epochs, the model with the best performance during training was saved. For the second to fifth batches, the network weight parameters of the best-performing model from the previous batch were used as the initial parameters. The model with the best performance during each batch was saved.

#### 2.5.3 Model Training With Data Augmentation

The data division and training method were the same as those for model training without data augmentation. First, the original database was randomly divided into 25 equal portions. Then, each small portion of data was analyzed to count the number of nodules in each category. And the images containing the categories that had fewer number were augmented to make the number of the categories increase.

The training method was consistent with that for model training without data augmentation, involving 5 batches of training with 80 epochs each. Except for the first batch, which used randomly initialized network weight parameters, the subsequent batches used the network weight parameters of the best-performing model from the previous batch.

## 3. Experimental Results

### 3.1 Quality of Junior Physicians’ Annotation of Thyroid Nodule Categories in Ultrasound Images

The overall accuracy of junior physicians’ annotation of thyroid nodule categories in ultrasound images was 76.9%. However, the accuracy varied greatly across categories. The accuracy for categories 2, 3, and 4A nodules was close to 80%, while that for categories 4B, 4C, and 5 nodules was less than 50%. Categories 4B, 4C, and 5 nodules were most frequently misclassified as category 4A. In a few images, junior physicians misclassified vascular, lymph node, and parathyroid structures within and around the thyroid gland as nodules.

Since categories 2, 3, and 4A nodules accounted for 95.7% of the total nodules, and categories 4B, 4C, and 5 nodules accounted for only 4.3%, the overall accuracy was 76.9%.

### 3.2 Model Training Results Using the Original Dataset

The average accuracy of the model trained using the original dataset was 55.1%, significantly lower than that of junior physicians.

### 3.3 Model Training Results Using the Augmented Data Set

Using the augmented data set, the first batch achieved an average accuracy of 49% for thyroid nodule classification in ultrasound images. The first batch’s training data set consisted of 1,360 original images and their augmented images, and the validation data set consisted of 340 original images and their augmented images. The fifth batch achieved an accuracy of 76.5%, very close to junior physicians’ accuracy of 76.9%.

By comparing Fig 8 and Fig 4, we found that junior physicians’ classification accuracy and that of the model trained on augmented data were very similar, but for categories 4A, 4B, 4C, and 5 nodules, the model’s accuracy was significantly higher than that of junior physicians. Junior physicians’ accuracy for these categories was less than 50%, while the model’s accuracy was around 80%, with the highest accuracy of 89% for category 4C nodules. For categories 2 and 3 nodules, the accuracy of both was very close.

**Fig 4:**
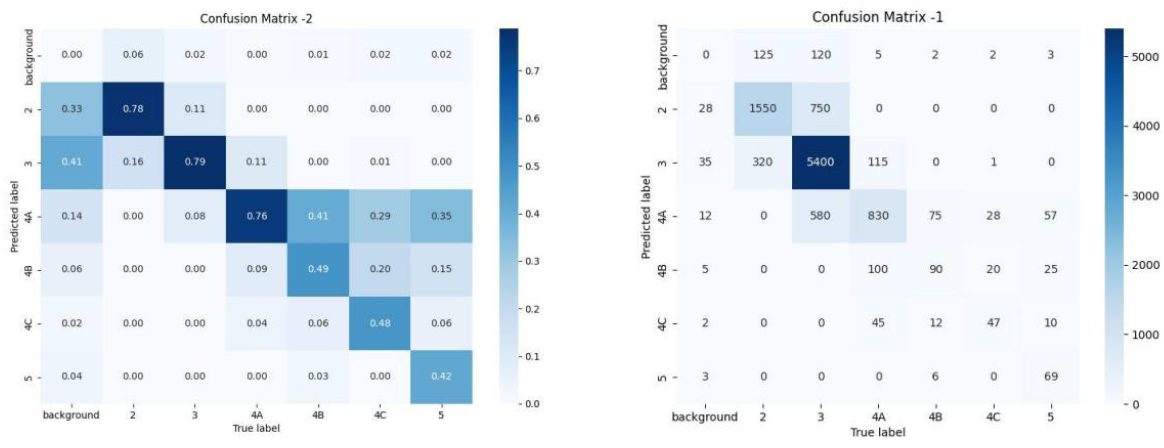
Confusion matrix of junior physicians’ annotation of thyroid nodule categories in ultrasound images. Fig 4 is a confusion matrix. The left half of the Fig is a proportional confusion matrix, and the right half is a quantitative confusion matrix. It shows the accuracy of each nodule category and the misclassification categories. In the confusion matrix, the horizontal axis represents the correct classification after senior physician review, and the vertical axis represents the classification by junior physicians. It can be seen that the accuracy for categories 2, 3, and 4A nodules was close to 80%, while that for categories 4B, 4C, and 5 nodules was less than 50%. Categories 4B, 4C, and 5 nodules were most frequently misclassified as category 4A.

## 4. Discussion

The cost of building a medical image database is very high. On one hand, the cost arises from the labor involved in annotation, as annotators must be licensed practitioners with experience, and the final annotations need to be reviewed and confirmed by senior doctors. On the other hand, the imbalance in the number of cases across categories poses a challenge, that some rare diseases having very few cases. Furthermore, images included in the database must meet additional requirements, such as being free of auxiliary markers like arrows, which are commonly found in workstation images. Therefore, most images in the database are newly collected, and collecting these case images, especially those of rare categories, can be a time-consuming process.

### 4.1

This study finds that step-by-step training the model during the data collection phase and using the trained model to pre-annotate new images is a feasible approach to reducing labor costs.

According to Fig 7, if the model trained with augmented data is used to pre-annotate new images, using the model trained on the first batch of data to annotate the second batch can reduce the annotation workload by a minimum of approximately 26.1% (when the model’s annotations and those of junior doctors are most inconsistent) and a maximum of approximately 49.2% (when they are most consistent). By the time the model trained on the first four batches of data is used to annotate the fifth batch, the workload can be reduced by at least 47.4% and up to 70.5%. The accuracy of the model trained on the first five batches of data is comparable to that of junior doctors, meaning that we can skip the pre-annotation by junior doctors and directly use the trained model to annotate images, thus saving 100% of the junior doctors’ workload.

### 4.2

There is a significant disparity in the number of cases across different medical image categories, with some categories having hundreds of times more cases than others. For instance, in the original database used in this study, the number of category 3 nodules is approximately 70 times that of category 4C nodules. Training the model directly with original data set results in poor performance, as evidenced by Figs 5 and 6, particularly for categories with fewer cases. However, through image data augmentation, as shown in Figs 7 and 8, the model’s training performance is significantly improved. Using a database augmented from just over 8,000 original images, the trained model achieves an average accuracy comparable to that of junior doctors’ annotations. For categories with fewer cases such as 4A, 4B, 4C, and 5, the model’s prediction accuracy can significantly surpass that of junior doctors.

**Fig 5:**
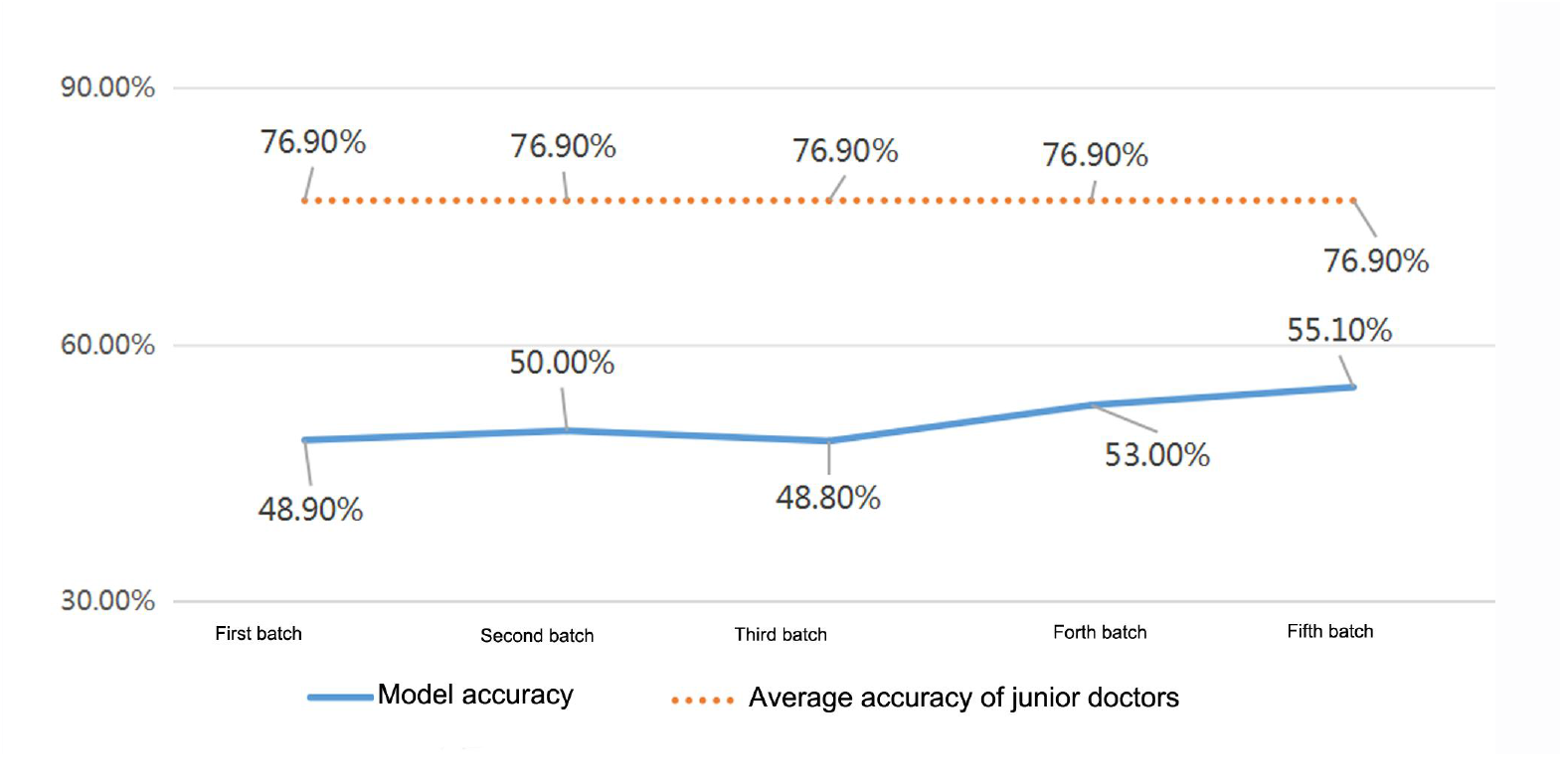
Comparison of nodule classification accuracy between junior physicians and the model trained using the original dataset. The orange dashed line in the Fig represents the average accuracy of junior physicians’ annotation of thyroid nodule categories in ultrasound images, and the blue line represents the average accuracy of the model trained using the original data set. It can be seen that junior physicians’ average accuracy was much higher than that of the model.

**Fig 6:**
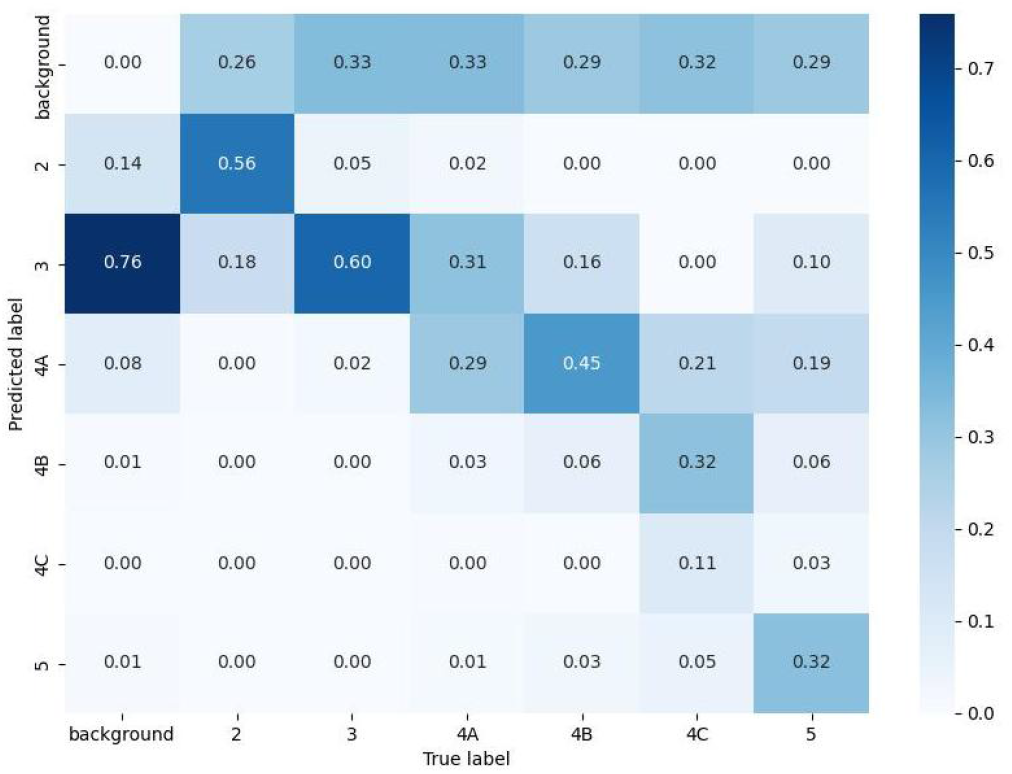
Confusion matrix of the model trained using the original dataset. Fig 6 is a proportional confusion matrix of the model trained using the original data set for the fifth batch. It can be seen that the highest classification accuracy was 60% for category 3 nodules, and the lowest was 6% for category 4B nodules, both significantly lower than junior physicians’ accuracy shown in Fig 4.

**Fig 7:**
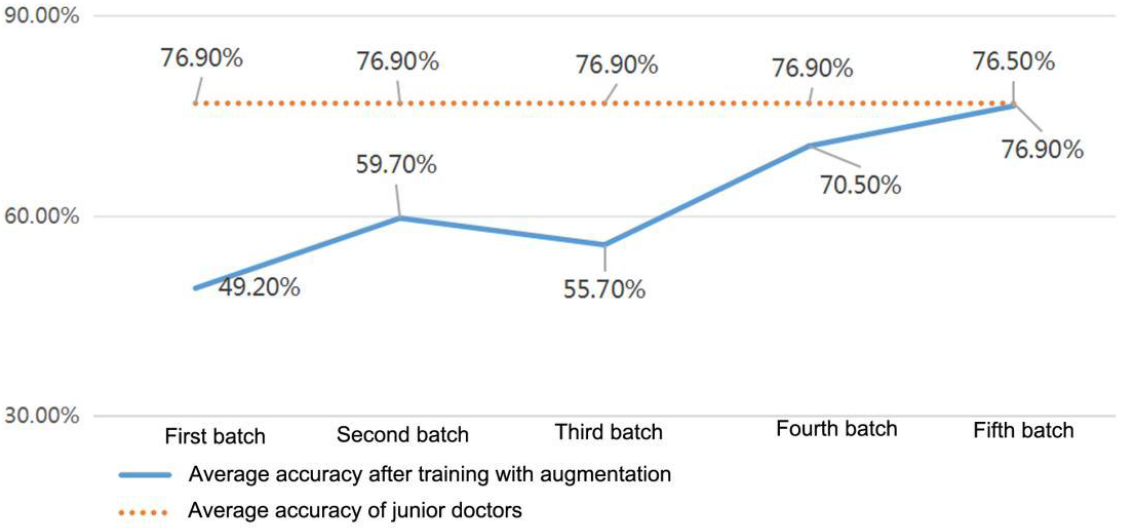
Comparison of Nodule Classification Accuracy in Ultrasound Images Between Junior Doctors and Models Trained with Augmented Data. The orange dashed line in the Fig represents the average accuracy of junior physicians’ annotation of thyroid nodule categories in ultrasound images, and the blue line represents the average accuracy of the model trained using the original data set. The average accuracy of nodule annotation in the augmented model has improved from 49% to 76.5%, reaching the accuracy level of junior doctors’ annotations.

**Fig 8:**
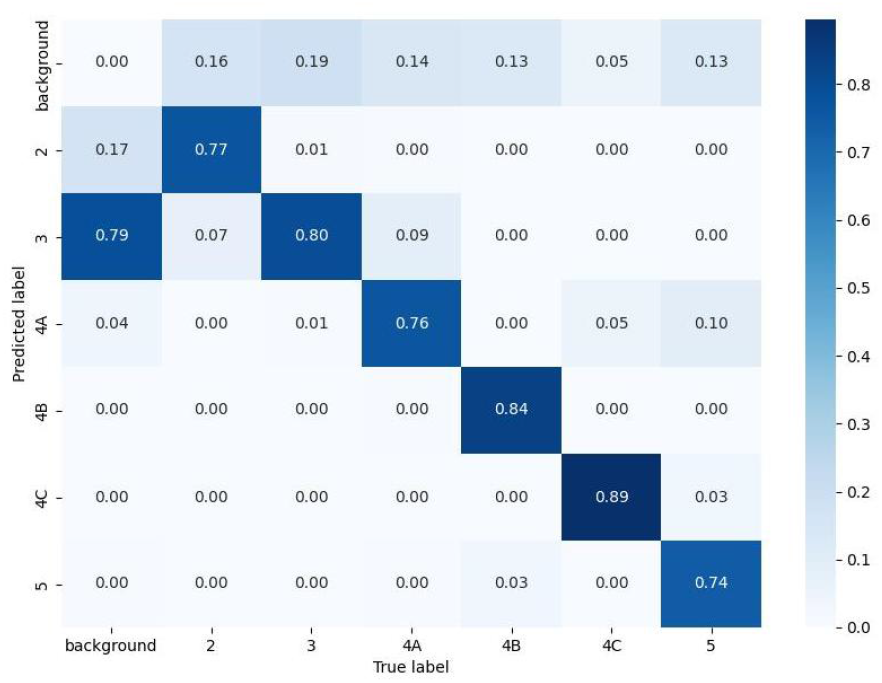
Confusion Matrix of Model Trained with Augmented Data for Thyroid Nodule Classification in Ultrasound Images. presents the confusion matrix for the classification of thyroid nodules in ultrasound images using the model trained with augmented data. It can be observed that the classification accuracy is lowest for category 5 nodules, at 74%, and highest for category 4C nodules, at 89%

**Fig 9:**
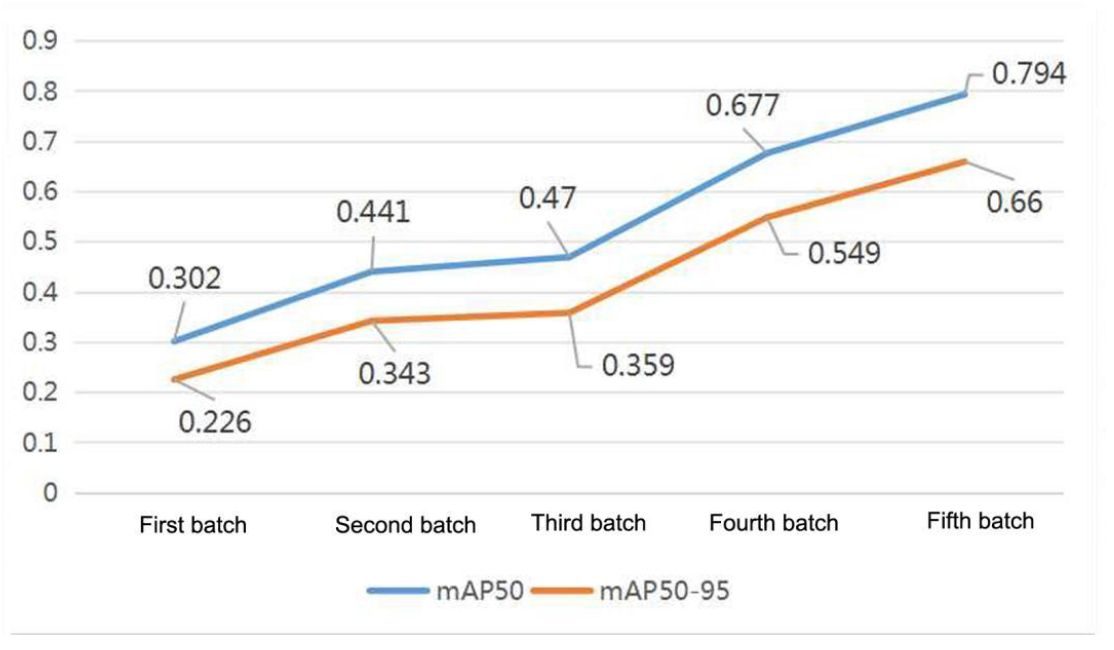
mAP50 and mAP50-95 of the Model Trained with Augmented Data. As shown in Fig 9, with the increase in training batches, both mAP50 and mAP50-95 continuously increase. After the fifth batch of training, the model’s mAP50 is 0.794, and mAP50-95 is 0.66.

### 4.3

Image classification tasks are widespread in medical imaging. This study, using thyroid nodule classification in ultrasound images as an example, has certain representativeness. Future research can explore similar tasks such as breast nodule classification, lymph node classification, and liver nodule classification in ultrasound images. Additionally, the images used in this study are high-quality with clear and typical nodule sections. In real-world scenarios, nodules need to be dynamically identified in video, and there may be interferences such as blurring, acoustic shadows, and atypical morphologies. The image augmentation methods used in this study may help improve the robustness of models in real-world applications. Future research will collect real-world images for validation.

### 4.4

Considering the small initial sample size for the step-by-step model training approach, k-fold cross-validation was not used to further optimize the model’s hyperparameters to prevent overfitting to the small sample data. For the same reason, each batch was trained for only 80 epochs.

## 5. Conclusion

Image data augmentation can reduce the impact of poor model training performance caused by imbalance across ultrasound image categories. Through step-by-step annotation, training, and using the step-by-step trained models to assist in image annotation and bounding box selection, the efficiency of pre-annotation by junior doctors can be improved. Ultimately, this approach can replace the pre-annotation work of junior doctors, significantly reducing the cost of data construction.

## Data Availability

The data is available at the URL: pan.baidu.com/s/1RpdpMUfnEobu6aSoPx6ckA?pwd=y00v

https://pan.baidu.com/s/15hYqvKUs-j5bhMAKsMfwvw?pwd=x94s

## Notes

### Competing Interest Statement

The authors have declared no competing interest.

### Funding Statement

Yes

